## Supplementary material for "Implication of Hepsin from primary tumor in the prognosis of colorectal cancer patients"

**Supplementary Table 1. Percentage of patients in the overall, localized and metastatic cohort with different intensities of Hepsin staining in each of the discrete clinical and histopathological categories recorded.** The p-values of Pearson's Chi-square correlations performed between hepsin and these variables are shown. *HPN*: Hepsin; *N (%)*: Number and percentage of patients; *ECOG*: Eastern Cooperative Oncology Group; *TNM II*: T3/4, N0, M0; *TNM III*: T1/2/3/4, N1/2, M0; *TNM* IV: T1/2/3/4, N0/1/2, M1; *T stage*: size and extension of the primary tumor; *N stage*: tumor involvement of nearby lymph nodes.

| **Overall cohort** | HPN staining intensity | | | p-value from Chi-square test |
|---|---|---|---|---|
| Clinical/histopathological variables correlated with HPN | Low | Medium | High | |
| Sex | | | | 0.819 |
| Men: N (%) | 39 (20.9) | 66 (35.3) | 82 (43.9) | |
| Women: N (%) | 22 (22.0) | 38 (38.0) | 40 (40.0) | |
| ECOG at diagnosis | | | | 0.736 |
| 0: N (%) | 23 (20.0) | 48 (41.7) | 44 (38.3) | |
| 1: N (%) | 27 (22.1) | 40 (32.8) | 55 (45.1) | |
| 2: N (%) | 5 (15.2) | 11 (33.3) | 17 (51.5) | |
| ≥3: N (%) | 1 (25.0) | 1 (25.0) | 2 (50.0) | |
| Primary tumor site | | | | 0.551 |
| Ascending colon: N (%) | 19 (22.9) | 31 (37.3) | 33 (39.8) | |
| Descending colon: N (%) | 1 (7.1) | 5 (35.7) | 8 (57.1) | |
| Transverse colon: N (%) | 3 (20.0) | 8 (53.3) | 4 (26.7) | |
| Sigmoid colon: N (%) | 12 (18.5) | 24 (36.9) | 29 (44.6) | |
| Rectal: N (%) | 26 (26.3) | 30 (30.3) | 43 (43.4) | |
| Number of chemotherapy lines | | | | 0.231 |
| 1: N (%) | 11 (21.2) | 20 (38.5) | 21 (40.4) | |
| 2: N (%) | 11 (28.2) | 10 (25.6) | 18 (46.2) | |
| 3: N (%) | 5 (14.3) | 17 (48.6) | 13 (37.1) | |
| 4: N (%) | 3 (20.0) | 4 (26.7) | 8 (53.3) | |
| ≥5: N (%) | 0 (0.0) | 9 (52.9) | 8 (47.1) | |
| RAS mutations | | | | 0.574 |
| Yes: N (%) | 17 (17.3) | 38 (38.8) | 43 (43.9) | |
| No: N (%) | 15 (23.1) | 21 (32.3) | 29 (44.6) | |
| BRAF mutations | | | | 0.579 |
| Yes: N (%) | 2 (33.3) | 1 (16.7) | 3 (50.0) | |
| No: N (%) | 27 (20.5) | 47 (35.6) | 58 (43.9) | |
| Microsatellite instability | | | | 0.172 |
| Yes: N (%) | 2 (13.3) | 3 (20.0) | 10 (66.7) | |
| No: N (%) | 26 (21.8) | 44 (37.0) | 49 (41.2) | |
| Histologic grade | | | | **<0.001** |
| Well differentiated: N (%) | 22 (17.3) | 55 (43.3) | 50 (39.4) | |
| Moderately differentiated: N (%) | 14 (16.5) | 26 (30.6) | 45 (52.9) | |

| | | | | |
|---|---|---|---|---|
| Poorly differentiated: N (%) | 13 (46.4) | 4 (14.3) | 11 (39.3) | |
| Lymphovascular invasion | | | | 0.656 |
| No: N (%) | 21 (21.9) | 35 (36.5) | 40 (41.7) | |
| Yes: N (%) | 21 (17.1) | 46 (37.4) | 56 (45.5) | |
| Perineural invasion | | | | 0.633 |
| No: N (%) | 31 (19.4) | 56 (35.0) | 73 (45.6) | |
| Yes: N (%) | 10 (17.5) | 24 (42.1) | 23 (40.4) | |
| Presence of tumor deposits distant to the primary tumor | | | | 0.197 |
| No: N (%) | 36 (20.6) | 62 (35.4) | 77 (44.0) | |
| Yes: N (%) | 5 (12.5) | 20 (50.0) | 15 (37.5) | |
| Tumor stages: TNM II vs III vs IV | | | | 0.727 |
| II: N (%) | 12 (21.8) | 19 (34.5) | 24 (43.6) | |
| III: N (%) | 14 (15.6) | 36 (40.0) | 40 (44.4) | |
| IV: N (%) | 27 (22.9) | 40 (33.9) | 51 (43.2) | |
| Tumor stages: TNM II-III vs IV | | | | 0.750 |
| II-III: N (%) | 34 (20.1) | 64 (37.9) | 71 (42.0) | |
| IV: N (%) | 27 (22.9) | 40 (33.9) | 51 (43.2) | |
| T stage | | | | 0.110 |
| T1: N (%) | 2 (25.0) | 2 (25.0) | 4 (50.0) | |
| T2: N (%) | 3 (17.6) | 3 (17.6) | 11 (64.7) | |
| T3: N (%) | 23 (16.1) | 53 (37.1) | 67 (46.9) | |
| T4: N (%) | 12 (25.0) | 23 (47.9) | 13 (27.1) | |
| N stage | | | | 0.611 |
| N0: N (%) | 16 (21.3) | 27 (36.0) | 32 (42.7) | |
| N1: N (%) | 16 (19.5) | 33 (40.2) | 33 (40.2) | |
| N2: N (%) | 7 (12.3) | 21 (36.8) | 29 (50.9) | |

| **Localized cohort** | HPN staining intensity | | | p-value from Chi-square test |
|---|---|---|---|---|
| Clinical/histopathological variables correlated with HPN | Low | Medium | High | |
| Sex | | | | 0.902 |
| Men: N (%) | 21 (19.3) | 41 (37.6) | 47 (43.1) | |
| Women: N (%) | 13 (21.7) | 23 (38.3) | 24 (40.0) | |
| ECOG at diagnosis | | | | 0.334 |
| 0: N (%) | 15 (17.6) | 38 (44.7) | 32 (37.6) | |
| 1: N (%) | 15 (22.4) | 21 (31.3) | 31 (46.3) | |
| 2: N (%) | 2 (18.2) | 2 (18.2) | 7 (63.6) | |
| ≥3: N (%) | 0 (0.0) | 1 (100.0) | 0 (0.0) | |
| Primary tumor site | | | | 0.513 |
| Ascending colon: N (%) | 12 (25.0) | 16 (33.3) | 20 (41.7) | |
| Descending colon: N (%) | 1 (11.1) | 3 (33.3) | 5 (55.6) | |
| Transverse colon: N (%) | 1 (10.0) | 6 (60.0) | 3 (30.0) | |
| Sigmoid colon: N (%) | 4 (11.4) | 16 (45.7) | 15 (42.9) | |
| Rectal: N (%) | 16 (25.0) | 21 (32.8) | 27 (42.2) | |

| | | | | |
|---|---|---|---|---|
| Number of chemotherapy lines | | | | 0.275 |
| 1: N (%) | 5 (22.7) | 6 (27.3) | 11 (50.0) | |
| 2: N (%) | 0 (0.0) | 5 (50.0) | 5 (50.0) | |
| 3: N (%) | 0 (0.0) | 6 (50.0) | 6 (50.0) | |
| 4: N (%) | 0 (0.0) | 1 (33.3) | 2 (66.6) | |
| ≥5: N (%) | 1 (25.0) | 3 (75.0) | 0 (0.0) | |
| RAS mutations | | | | 0.381 |
| Yes: N (%) | 2 (5.7) | 16 (45.7) | 17 (48.6) | |
| No: N (%) | 4 (16.7) | 9 (37.5) | 11 (45.8) | |
| BRAF mutations | | | | 0.168 |
| Yes: N (%) | 1 (50.0) | 0 (0.0) | 1 (50.0) | |
| No: N (%) | 5 (9.8) | 21 (41.2) | 25 (49.0) | |
| Microsatellite instability | | | | 0.218 |
| Yes: N (%) | 2 (15.4) | 2 (15.4) | 9 (69.2) | |
| No: N (%) | 13 (17.1) | 29 (38.2) | 34 (44.7) | |
| Histologic grade | | | | 0.057 |
| Well differentiated: N (%) | 17 (18.9) | 33 (36.7) | 40 (44.4) | |
| Moderately differentiated: N (%) | 10 (18.9) | 23 (43.4) | 20 (37.7) | |
| Poorly differentiated: N (%) | 5 (50.0) | 0 (0.0) | 5 (50.0) | |
| Lymphovascular invasion | | | | 0.626 |
| No: N (%) | 16 (21.3) | 28 (37.3) | 31 (41.3) | |
| Yes: N (%) | 12 (15.8) | 28 (36.8) | 36 (47.4) | |
| Perineural invasion | | | | 0.653 |
| No: N (%) | 23 (19.7) | 44 (37.6) | 50 (42.7) | |
| Yes: N (%) | 5 (15.2) | 11 (33.3) | 17 (51.5) | |
| Presence of tumor deposits distant to the primary tumor | | | | 0.156 |
| No: N (%) | 26 (20.5) | 46 (36.2) | 55 (43.3) | |
| Yes: N (%) | 2 (10.0) | 9 (45.0) | 9 (45.0) | |
| Tumor stages: TNM II vs III | | | | 0.601 |
| II: N (%) | 12 (21.8) | 19 (34.5) | 24 (43.6) | |
| III: N (%) | 14 (15.6) | 36 (40.0) | 40 (44.4) | |
| T stage | | | | 0.256 |
| T1: N (%) | 2 (40.0) | 1 (20.0) | 2 (40.0) | |
| T2: N (%) | 3 (20.0) | 3 (20.0) | 9 (60.0) | |
| T3: N (%) | 17 (16.0) | 40 (37.7) | 49 (46.2) | |
| T4: N (%) | 5 (23.8) | 11 (52.4) | 5 (23.8) | |
| N stage | | | | 0.357 |
| N0: N (%) | 12 (21.8) | 19 (34.5) | 24 (43.6) | |
| N1: N (%) | 12 (20.3) | 24 (40.7) | 23 (39.0) | |
| N2: N (%) | 2 (6.5) | 12 (38.7) | 17 (54.8) | |
| **Metastatic cohort** | HPN staining intensity | | | p-value from Chi-square test |
| Clinical/histopathological variables correlated with HPN | Low | Medium | High | |

| | | | | |
|---|---|---|---|---|
| Sex | | | | 0.827 |
| Men: N (%) | 18 (23.1) | 25 (32.1) | 35 (44.9) | |
| Women: N (%) | 9 (22.5) | 15 (37.5) | 16 (40.0) | |
| ECOG at diagnosis | | | | 0.814 |
| 0: N (%) | 8 (26.7) | 10 (33.3) | 12 (40.0) | |
| 1: N (%) | 12 (21.8) | 19 (34.5) | 24 (43.6) | |
| 2: N (%) | 3 (13.6) | 9 (40.9) | 10 (45.5) | |
| ≥3: N (%) | 1 (33.3) | 0 (0.0) | 2 (66.7) | |
| Primary tumor site | | | | 0.645 |
| Ascending colon: N (%) | 7 (20.0) | 15 (42.9) | 13 (37.1) | |
| Descending colon: N (%) | 0 (0.0) | 2 (40.0) | 3 (60.0) | |
| Transverse colon: N (%) | 2 (40.0) | 2 (40.0) | 1 (20.0) | |
| Sigmoid colon: N (%) | 8 (26.7) | 8 (26.7) | 14 (46.7) | |
| Rectal: N (%) | 10 (28.6) | 9 (25.7) | 16 (45.7) | |
| Liver metastasis at diagnosis | | | | 0.810 |
| Yes: N (%) | 20 (23.8) | 27 (32.1) | 37 (44.0) | |
| No: N (%) | 7 (20.6) | 13 (38.2) | 14 (41.2) | |
| Peritoneal metastasis at diagnosis | | | | 0.110 |
| Yes: N (%) | 11 (34.4) | 7 (21.9) | 14 (43.8) | |
| No: N (%) | 16 (18.6) | 33 (38.4) | 37 (43.0) | |
| Lung metastasis at diagnosis | | | | 0.245 |
| Yes: N (%) | 10 (32.3) | 11 (35.5) | 10 (32.3) | |
| No: N (%) | 17 (19.5) | 29 (33.3) | 41 (47.1) | |
| >1 metastasis site at diagnosis | | | | **0.023** |
| Yes: N (%) | 16 (34.8) | 16 (34.8) | 14 (30.4) | |
| No: N (%) | 11 (15.3) | 24 (33.3) | 37 (51.4) | |
| Number of chemotherapy lines | | | | 0.106 |
| 1: N (%) | 6 (20.0) | 14 (46.7) | 10 (33.3) | |
| 2: N (%) | 11 (37.9) | 5 (17.2) | 13 (44.8) | |
| 3: N (%) | 5 (21.7) | 11 (47.8) | 7 (30.4) | |
| 4: N (%) | 3 (25.0) | 3 (25.0) | 6 (50.0) | |
| ≥5: N (%) | 0 (0.0) | 6 (46.2) | 7 (53.8) | |
| RAS mutations | | | | 0.829 |
| Yes: N (%) | 15 (23.8) | 22 (34.9) | 26 (41.3) | |
| No: N (%) | 11 (26.8) | 12 (29.3) | 18 (43.9) | |
| BRAF mutations | | | | 0.929 |
| Yes: N (%) | 1 (25.0) | 1 (25.0) | 2 (50.0) | |
| No: N (%) | 22 (27.2) | 26 (32.1) | 33 (40.7) | |
| Microsatellite instability | | | | 0.654 |
| Yes: N (%) | 0 (0.0) | 1 (50.0) | 1 (50.0) | |
| No: N (%) | 13 (30.2) | 15 (34.9) | 15 (34.9) | |
| Histologic grade | | | | **<0.001** |
| Well differentiated: N (%) | 5 (13.5) | 22 (59.5) | 10 (27.0) | |
| Moderately differentiated: N (%) | 4 (12.5) | 3 (9.4) | 25 (78.1) | |

| | | | | |
|---|---|---|---|---|
| Poorly differentiated: N (%) | 8 (44.4) | 4 (22.2) | 6 (33.3) | |
| Lymphovascular invasion | | | | 0.626 |
| No: N (%) | 16 (21.3) | 28 (37.3) | 31 (41.3) | |
| Yes: N (%) | 12 (15.8) | 28 (36.8) | 36 (47.4) | |
| Perineural invasion | | | | 0.653 |
| No: N (%) | 23 (19.7) | 44 (37.6) | 50 (42.7) | |
| Yes: N (%) | 5 (15.2) | 11 (33.3) | 17 (51.5) | |
| Presence of tumor deposits distant to the primary tumor | | | | 0.156 |
| No: N (%) | 26 (20.5) | 46 (36.2) | 55 (43.3) | |
| Yes: N (%) | 2 (10.0) | 9 (45.0) | 9 (45.0) | |
| T stage | | | | 0.399 |
| T1: N (%) | 0 (0.0) | 1 (33.3) | 2 (66.6) | |
| T2: N (%) | 0 (0.0) | 0 (0.0) | 2 (100.0) | |
| T3: N (%) | 6 (16.2) | 13 (35.1) | 18 (48.6) | |
| T4: N (%) | 7 (25.9) | 12 (44.4) | 8 (29.6) | |
| N stage | | | | 0.993 |
| N0: N (%) | 4 (20.0) | 8 (40.0) | 8 (40.0) | |
| N1: N (%) | 4 (17.4) | 9 (39.1) | 10 (43.5) | |
| N2: N (%) | 5 (19.2) | 9 (34.6) | 12 (46.2) | |

**Supplementary Table 2. Correlations between Hepsin staining intensity and time-to-event variables.** The results of these correlations are shown for the overall cohort and localized and metastatic patients separately. *HPN*: Hepsin; *N*: number of patients for each hepsin staining intensity; *CI*: confidence interval; *LL*: lower limit; *UL*: upper limit; *tft*: log rank test for trends; *Stat*: statistical value; *df*: degrees of freedom. *Follow-up limit reached.

**Overall cohort**

| HPN staining intensity | N | Death events | 5-year overall survival (%) | Standard error (%) | LL CI 95% (%) | UL CI 95% (%) | p-value tft |
|---|---|---|---|---|---|---|---|
| High | 121 | 60 | 41 | 6 | 31 | 55 | 0.40 |
| Medium | 104 | 52 | 59 | 6 | 49 | 71 | |
| Low | 61 | 29 | 55 | 7 | 43 | 71 | |

| HPN staining intensity | N | Thrombosis events | 5-year cumulative incidence of thrombosis (%) | Standard error (%) | LL CI 95% (%) | UL CI 95% (%) | p-value tft | Gray´s test | | |
|---|---|---|---|---|---|---|---|---|---|---|
| | | | | | | | | Stat | df | p-value |
| High | 121 | 31 | 30 | 5 | 20 | 38 | 0.61 | 2.60 | 2 | 0.27 |
| Medium | 104 | 24 | 26 | 5 | 15 | 35 | | | | |
| Low | 61 | 10 | 15 | 5 | 5 | 24 | | | | |

**Localized cohort**

| HPN staining intensity | N | Death events | 5-year overall survival (%) | Standard error (%) | LL CI 95% (%) | UL CI 95% (%) | p-value tft |
|---|---|---|---|---|---|---|---|
| High | 71 | 16 | 63 | 9 | 48 | 83 | 0.67 |
| Medium | 64 | 22 | 70 | 7 | 58 | 86 | |
| Low | 34 | 8 | 81 | 8 | 67 | 99 | |

| HPN staining intensity | N | Metastatic relapse events | 5-year disease-free survival (%) | Standard error (%) | LL CI 95% (%) | UL CI 95% (%) | p-value tft |
|---|---|---|---|---|---|---|---|
| High | 71 | 26 | 51 | 8 | 37 | 71 | 0.16 |
| Medium | 64 | 22 | 59 | 7 | 46 | 75 | |
| Low | 34 | 6 | 73 | 10 | 56 | 96 | |

| HPN staining intensity | N | Thrombosis events | 5-year cumulative incidence of thrombosis (%) | Standard error (%) | LL CI 95% (%) | UL CI 95% (%) | p-value tft | Gray´s test | | |
|---|---|---|---|---|---|---|---|---|---|---|
| | | | | | | | | Stat | df | p-value |
| High | 71 | 16 | 23 | 5 | 12 | 33 | | | | |
| Medium | 64 | 13 | 22 | 6 | 9 | 33 | **0.038** | 19.38 | 2 | **0.009** |
| Low | 34 | 1 | 0 | 0 | 0 | 0 | | | | |

**Metastatic cohort**

| HPN staining intensity | N | Death events | 5-year overall survival (%) | Standard error (%) | LL CI 95% (%) | UL CI 95% (%) | p-value tft |
|---|---|---|---|---|---|---|---|
| High | 50 | 44 | 18 | 6 | 9 | 34 | |
| Medium | 40 | 30 | 41 | 8 | 28 | 61 | 0.21 |
| Low | 27 | 21 | 24 | 8 | 12 | 48 | |

| HPN staining intensity | N | Progression events | 4-year* progression-free survival (%) | Standard error (%) | LL CI 95% (%) | UL CI 95% (%) | p-value tft |
|---|---|---|---|---|---|---|---|
| High | 44 | 43 | 5 | 3 | 1 | 18 | |
| Medium | 38 | 35 | 11 | 5 | 4 | 27 | 0.20 |
| Low | 25 | 24 | 4 | 4 | 1 | 27 | |

| HPN staining intensity | N | Thrombosis events | 5-year cumulative incidence of thrombosis (%) | Standard error (%) | LL CI 95% (%) | UL CI 95% (%) | p-value tft | Gray´s test | | |
|---|---|---|---|---|---|---|---|---|---|---|
| | | | | | | | | Stat | df | p-value |
| High | 50 | 15 | 43 | 9 | 22 | 58 | | | | |
| Medium | 40 | 11 | 31 | 9 | 11 | 46 | 0.23 | 0.83 | 2 | 0.66 |
| Low | 27 | 9 | 35 | 11 | 13 | 52 | | | | |

**Supplementary Table 3. Correlations between Hepsin staining intensity and continuous clinical-histopathological variables.** The results of these correlations are shown for the overall cohort and localized and metastatic patients separately. *HPN*: Hepsin; *CEA*: carcinoembryonic antigen; *LDH*: lactate dehydrogenase.

| **Overall cohort** | HPN staining intensity | | | p-value from Kruskal-Wallis test |
|---|---|---|---|---|
| Clinical/histopathological variables correlated with HPN | Low | Medium | High | |
| Age at diagnosis (years): Mean; standard error of mean | 62.5; 1.8 | 62.9; 1.2 | 66.1; 0.9 | 0.176 |
| CEA (ng/ml): Mean; standard error of mean | 49.1; 20.4 | 30.7; 8.9 | 22.1; 4.7 | 0.334 |
| Leukocytes (x10^3/µl): Mean; standard error of mean | 8.0; 0.3 | 8.6; 0.4 | 8.3; 0.3 | 0.573 |
| Platelets (x10^3/µl): Mean; standard error of mean | 307.0; 17.7 | 281.0; 12.7 | 278.0; 10.9 | 0.368 |
| LDH (U/l): Mean; standard error of mean | 420.0; 76.3 | 497.0; 63.1 | 578.0; 107.0 | 0.828 |
| Number of tumor deposits distant to the primary tumor: Mean; standard error of mean | 1.0; 0.7 | 0.8; 0.2 | 0.4; 0.1 | 0.508 |
| **Localized cohort** | HPN staining intensity | | | p-value from Kruskal-Wallis test |
| Clinical/histopathological variables correlated with HPN | Low | Medium | High | |
| Age at diagnosis (years): Mean; standard error of mean | 60.9; 2.3 | 62.3; 1.4 | 65.4; 1.1 | 0.223 |
| CEA (ng/ml): Mean; standard error of mean | 5.8; 2.7 | 6.9; 1.9 | 6.6; 1.7 | 0.315 |
| Leukocytes (x10^3/µl): Mean; standard error of mean | 7.5; 0.4 | 7.8; 0.4 | 7.7; 0.4 | 0.964 |
| Platelets (x10^3/µl): Mean; standard error of mean | 304.0; 27.8 | 279.0; 14.7 | 271.0; 14.6 | 0.684 |
| LDH (U/l): Mean; standard error of mean | 310.0; 35.1 | 338.0; 18.1 | 330.0; 17.4 | 0.809 |
| Number of tumor deposits distant to the primary tumor: Mean; standard error of mean | 1.1; 1.1 | 0.5; 0.2 | 0.3; 0.1 | 0.664 |
| **Metastatic cohort** | HPN staining intensity | | | p-value from Kruskal-Wallis test |
| Clinical/histopathological variables correlated with HPN | Low | Medium | High | |
| Age at diagnosis (years): Mean; standard error of mean | 64.5; 2.7 | 63.9; 2.2 | 66.9; 1.5 | 0.727 |

| | | | | |
|---|---|---|---|---|
| CEA (ng/ml): Mean; standard error of mean | 106.0; 44.4 | 72.5; 22.5 | 48.3; 11.0 | 0.614 |
| Leukocytes (x10^3/µl): Mean; standard error of mean | 8.7; 0.5 | 9.9; 0.6 | 9.1; 0.6 | 0.481 |
| Platelets (x10^3/µl): Mean; standard error of mean | 311.0; 19.2 | 284.0; 23.2 | 289.0; 16.2 | 0.527 |
| LDH (U/l): Mean; standard error of mean | 496.0; 124.0 | 685.0; 122.0 | 939.0; 231.0 | 0.532 |
| Number of tumor deposits distant to the primary tumor: Mean; standard error of mean | 0.8; 0.4 | 1.4; 0.5 | 0.5; 0.2 | 0.417 |

**Supplementary Table 4. Clinical scenario of localized patients who suffered thrombosis during follow-up.** For each patient, a series of characteristics potentially underlying the origin of the thrombotic event are shown. *ID*: identifier; *PE*: pulmonary embolism; *DVT*: deep vein thrombosis; *NOS*: not otherwise specified; *NA*: not applicable; *CAPOX*: Capecitabine+Oxaliplatin; *FUOX*: Fluorouracil+Oxaliplatin; *5FU*: 5-Fluorouracil; *FOLFIRI*: Folinic acid+Fluorouracil+Irinotecan; *SNG+NTP:* Nasogastric tube+parenteral nutrition.

| Patient ID | Location of thrombosis | Months until thrombosis since tumor diagnosis | Patient suffers metastasis relapse before thrombosis? | Months between relapse and thrombosis | Hemostasis-related background before tumor diagnosis | Chemotherapy treatment before thrombosis | Months between chemotherapy and thrombosis | Particular incidents of each patient |
|---|---|---|---|---|---|---|---|---|
| 1 | Lower extremity, NOS | 11.27 | No | NA | Arterial hypertension/ dyslipidemia/ supraventricular tachycardia | Adjuvant CAPOX | 8.83 | At diagnosis of the colorectal tumor, the patient also had a spiculated nodule in the left upper lobe (it could be a parenchymal scar vs a primary neoplasm) that persisted in August 2018, four months before thrombosis |
| 2 | DVT, NOS | 20.03 | No | NA | NA | NA | NA | NA |
| 3 | Head & neck | 26.40 | No | NA | NA | NA | NA | Patient was admitted to the hospital for ileostomy closure with |

| | | | | | | | | |
|---|---|---|---|---|---|---|---|---|
| | | | | | | | | transit reconstruction when he had the thrombosis |
| 4 | Lower extremity, NOS | 6.47 | No | NA | NA | Adjuvant Capecitabine | 3.47 | NA |
| 5 | Splanchnic | 4.90 | No | NA | Dyslipidemia | Adjuvant Capecitabine | 3.87 | NA |
| 6 | Femoral + PE | 2.03 | No | NA | NA | Neoadjuvant radiotherapy + Capecitabine | 1.20 | At the start of neoadjuvant treatment, patient suffered polymicrobial right sternoclavicular septic arthritis, with previous trauma in the affected area |
| 7 | PE, NOS | 0.47 | No | NA | NA | NA | NA | NA |
| 8 | Splanchnic | 8.77 | No | NA | NA | Adjuvant CAPOX | 2.67 | NA |
| 9 | Femoral | 6.87 | No | NA | Arterial hypertension | Adjuvant CAPOX | 3.97 | NA |
| 10 | PE, NOS | 2.10 | No | NA | Arterial hypertension/ overweight | Adjuvant CAPOX | 0.90 | Two months before thrombosis, a right hemicolectomy was performed, and after the operation, there was a prolonged ileus that was resolved with |

| | | | | | | | | |
|---|---|---|---|---|---|---|---|---|
| | | | | | | | | SNG+NTP treatment |
| 11 | PE, NOS | 5.17 | No | NA | NA | Adjuvant CAPOX | 3.13 | NA |
| 12 | Calf vein | 2.00 | No | NA | Arterial hypertension | Adjuvant CAPOX | 1.00 | NA |
| 13 | Femoral | 4.03 | No | NA | NA | Adjuvant CAPOX | 3.03 | NA |
| 14 | PE, NOS | 9.10 | No | NA | Dyslipidemia | Adjuvant CAPOX+FUOX | 6.50 | NA |
| 15 | Lower extremity, NOS | 2.13 | No | NA | NA | Adjuvant Capecitabine | 0.47 | NA |
| 16 | Catheter-related + PE | 0.53 | No | NA | NA | NA | NA | Patient had thrombosis the same month he had rectal surgery |
| 17 | PE, NOS | 4.07 | Yes | 2.30 | Arterial hypertension/ non-insulinized type-II diabetes mellitus | Oxaliplatin+Xeloda | 2.13 | One month before thrombosis, it was detected a doubtful uterine myoma respecting its status as a primary or metastatic tumor |
| 18 | Femoral | 33.43 | Yes | 3.03 | Arterial hypertension/ dyslipidemia/ type-II diabetes mellitus/ dyspnea on exertion | Capecitabine+Avastin | 2.47 | NA |
| 19 | Femoral | 47.20 | Yes | 24.30 | NA | 5FU+Avastin | 5.57 | NA |

| | | | | | | | | |
|---|---|---|---|---|---|---|---|---|
| 20 | PE, NOS | 35.10 | Yes | 9.23 | Dyslipidemia/ ischemic heart disease | Oxaliplatin+5FU+Bevacizumab | 11.40 | NA |
| 21 | PE, NOS | 12.37 | Yes | 0.10 | Dyslipidemia/ chronic arthritis in lower extremities | NA | NA | Patient was admitted to the hospital when he suffered thrombosis |
| 22 | Lower extremity, NOS | 22.33 | Yes | 3.00 | NA | Irinotecan+Cetuximab | 3.07 | NA |
| 23 | Splanchnic | 9.77 | Yes | 1.70 | Patient walks with help but with difficulty due to shortening of the right leg | NA | NA | NA |
| 24 | Head & neck | 114.90 | Yes | 58.40 | Arterial hypertension/ generalized osteoarthritis | NA | NA | The day when thrombosis was detected, patient had previously fallen twice at home |
| 25 | PE, NOS | 18.67 | Yes | 8.50 | NA | FOLFIRI+Avastin/Capecitabine+Avastin | 8.53 | NA |
| 26 | Splanchnic | 62.17 | Yes | 14.47 | Arterial hypertension/ dyslipidemia/ embolic cardiovascular accident | FOLFIRI | 8.43 | NA |
| 27 | Lower extremity, NOS | 28.03 | Yes | 11.80 | NA | FOLFIRI+Aflibercept | 6.73 | It was a patient treated with low molecular weight heparin who stopped taking it due to episodes of |

| | | | | | | | | |
|---|---|---|---|---|---|---|---|---|
| | | | | | | | | peristomal bleeding prior to thrombosis |
| 28 | Splanchnic | 3.60 | Yes | 0.27 | Thoracic aortic aneurysm/ knee osteoarthritis | NA | NA | NA |
| 29 | PE, NOS | 23.37 | Yes | 8.10 | Arterial hypertension/ polycythemia vera | NA | NA | NA |
| 30 | Splanchnic | 63.77 | Yes | 49.60 | Arterial hypertension/ dyslipidemia | Oxaliplatin+5FU+Bevacizumab | 36.97 | Postoperative thrombosis |

**Supplementary Table 5. Multivariable models for overall survival, progression-free survival and cumulative incidence of thrombosis in metastatic patients at diagnosis.** The risk of death is calculated according to Hepsin staining intensity, adjusting this calculation by adding different covariates to the model. The risk of progression is calculated according to Hepsin staining intensity, adjusting this calculation by adding different covariates to the model. The risk of thrombosis is calculated according to Hepsin staining intensity, adjusting this calculation by adding different covariates to the model. *OS*: overall survival; *Coef*: Cox regression coefficient; *exp (coef)*: hazard ratio; *CI*: confidence interval; *LL*: lower limit; *UL*: upper limit; *se (coef)*: standard error of Cox regression coefficient; *p*: p-value; *HPN*: Hepsin; *ECOG*: Eastern Cooperative Oncology Group; *PFS*: progression-free survival after the first line of chemotherapy.

| **Multivariate Cox regression for OS** | | | | | | |
|---|---|---|---|---|---|---|
| **Regressions** | **Coef** | **exp(coef)** | **LL CI 95% of exp(coef)** | **UL CI 95% of exp(coef)** | **se(coef)** | **p** |
| HPN Medium | 0.11 | 1.11 | 0.64 | 1.95 | 0.29 | 0.71 |
| HPN High | -0.05 | 0.95 | 0.50 | 1.80 | 0.33 | 0.87 |
| HPN Low (reference) | - | - | - | - | - | - |
| Moderately differentiated histological grade | 0.27 | 1.31 | 0.76 | 2.26 | 0.28 | 0.34 |
| Poorly differentiated histological grade | 1.28 | 3.59 | 1.88 | 6.83 | 0.33 | **0.0001** |
| Well differentiated histological grade (reference) | - | - | - | - | - | - |
| ECOG 1 | 0.33 | 1.39 | 0.81 | 2.38 | 0.27 | 0.23 |
| ECOG 2 | 0.84 | 2.31 | 1.22 | 4.38 | 0.33 | **0.01** |
| ECOG ≥3 | 2.50 | 12.23 | 3.07 | 48.70 | 0.70 | **0.0004** |
| ECOG 0 (reference) | - | - | - | - | - | - |
| >1 metastasis site | -0.28 | 0.76 | 0.40 | 1.42 | 0.32 | 0.39 |
| 1 metastasis site (reference) | - | - | - | - | - | - |

| | | | | | | |
|---|---|---|---|---|---|---|
| Peritoneal involvement | 0.13 | 1.14 | 0.57 | 2.25 | 0.35 | 0.71 |
| No peritoneal involvement (reference) | - | - | - | - | - | - |
| Lung involvement | 0.21 | 1.23 | 0.65 | 2.31 | 0.32 | 0.52 |
| No lung involvement (reference) | - | - | - | - | - | - |
| Liver involvement | 0.16 | 1.17 | 0.61 | 2.25 | 0.34 | 0.64 |
| No liver involvement (reference) | - | - | - | - | - | - |

| **Multivariate Cox regression for PFS** | | | | | | |
|---|---|---|---|---|---|---|
| **Regressions** | **Coef** | **exp(coef)** | **LL CI 95% of exp(coef)** | **UL CI 95% of exp(coef)** | **se(coef)** | **p** |
| HPN Medium | 0.14 | 1.15 | 0.64 | 2.04 | 0.29 | 0.64 |
| HPN High | 0.05 | 1.05 | 0.59 | 1.89 | 0.30 | 0.86 |
| HPN Low (reference) | - | - | - | - | - | - |
| Moderately differentiated histological grade | 0.47 | 1.60 | 0.94 | 2.70 | 0.27 | 0.08 |
| Poorly differentiated histological grade | 1.07 | 2.91 | 1.62 | 5.25 | 0.30 | **0.0004** |
| Well differentiated histological grade (reference) | - | - | - | - | - | - |
| ECOG 1 | -0.21 | 0.81 | 0.50 | 1.32 | 0.25 | 0.39 |
| ECOG 2 | 0.34 | 1.41 | 0.75 | 2.66 | 0.32 | 0.29 |
| ECOG ≥3 | 0.94 | 2.56 | 0.65 | 10.12 | 0.70 | 0.18 |
| ECOG 0 (reference) | - | - | - | - | - | - |
| >1 metastasis site | -0.16 | 0.85 | 0.45 | 1.64 | 0.33 | 0.63 |
| 1 metastasis site (reference) | - | - | - | - | - | - |

| | | | | | | |
|---|---|---|---|---|---|---|
| Peritoneal involvement | 0.31 | 1.36 | 0.62 | 3.00 | 0.40 | 0.45 |
| No peritoneal involvement (reference) | - | - | - | - | - | - |
| Lung involvement | 0.20 | 1.22 | 0.60 | 2.50 | 0.36 | 0.58 |
| No lung involvement (reference) | - | - | - | - | - | - |
| Liver involvement | 0.07 | 1.07 | 0.50 | 2.28 | 0.39 | 0.86 |
| No liver involvement (reference) | - | - | - | - | - | - |

| **Multivariate Fine&Gray regression for cumulative incidence of thrombosis** | | | | | | |
|---|---|---|---|---|---|---|
| **Regressions** | **Coef** | **exp(coef)** | **LL CI 95% of exp(coef)** | **UL CI 95% of exp(coef)** | **se(coef)** | **p** |
| HPN Medium | -0.15 | 0.86 | 0.30 | 2.47 | 0.54 | 0.78 |
| HPN High | -0.08 | 0.92 | 0.37 | 2.33 | 0.47 | 0.87 |
| HPN Low (reference) | - | - | - | - | - | - |
| Moderately differentiated histological grade | -0.18 | 0.84 | 0.34 | 2.05 | 0.46 | 0.69 |
| Poorly differentiated histological grade | 1.05 | 2.85 | 1.11 | 7.33 | 0.48 | **0.029** |
| Well differentiated histological grade (reference) | - | - | - | - | - | - |
| Lymphovascular invasion | 0.04 | 1.04 | 0.47 | 2.30 | 0.41 | 0.93 |
| Absent lymphovascular invasion (reference) | - | - | - | - | - | - |
| >1 metastasis site | -0.56 | 0.57 | 0.20 | 1.67 | 0.55 | 0.31 |
| 1 metastasis site (reference) | - | - | - | - | - | - |
| Peritoneal involvement | -0.32 | 0.73 | 0.25 | 2.12 | 0.55 | 0.56 |
| No peritoneal involvement (reference) | - | - | - | - | - | - |
| Lung involvement | 0.74 | 2.09 | 0.63 | 6.92 | 0.61 | 0.23 |
| No lung involvement (reference) | - | - | - | - | - | - |

| | | | | | | |
|---|---|---|---|---|---|---|
| Liver involvement | 0.24 | 1.27 | 0.44 | 3.68 | 0.54 | 0.66 |
| No liver involvement (reference) | - | - | - | - | - | - |

**Supplementary Table 6. Correlations between Hepsin staining intensity and time-to-event variables in metastatic patients with mutated RAS.** Upper table shows correlations based on the Kaplan-Meier estimator and the Log-rank for trends and Gray tests. Lower tables show Cox and Fine&Gray multivariate models. *HPN*: Hepsin; *N*: number of patients for each hepsin level; *CI*: confidence interval; *LL*: lower limit; *UL*: upper limit; *tft*: Log rank test for trends; *Stat*: statistical value; *df*: degrees of freedom; *OS*: overall survival; *Coef*: Cox regression coefficient; *exp (coef)*: hazard ratio; *se (coef)*: standard error of Cox regression coefficient; *p*: p-value; *ECOG*: Eastern Cooperative Oncology Group. *PFS*: progression-free survival after first-line chemotherapy. *Follow-up limit reached.

| HPN staining intensity | N | Death events | 5-year overall survival (%) | Standard error (%) | LL CI 95% (%) | UL CI 95% (%) | p-value tft |
|---|---|---|---|---|---|---|---|
| High | 26 | 25 | 5 | 5 | 1 | 31 | 0.09 |
| Medium | 22 | 16 | 39 | 11 | 23 | 67 | |
| Low | 15 | 11 | 23 | 11 | 9 | 61 | |

| HPN staining intensity | N | Progression events | 4-year* progression-free survival (%) | Standard error (%) | LL CI 95% (%) | UL CI 95% (%) | p-value tft |
|---|---|---|---|---|---|---|---|
| High | 23 | 22 | 9 | 6 | 2 | 33 | 0.42 |
| Medium | 22 | 20 | 9 | 6 | 2 | 34 | |
| Low | 15 | 14 | 7 | 6 | 1 | 44 | |

| HPN staining intensity | N | Thrombosis events | 4-year* cumulative incidence of thrombosis (%) | Standard error (%) | LL CI 95% (%) | UL CI 95% (%) | p-value tft | Gray´s test | | |
|---|---|---|---|---|---|---|---|---|---|---|
| | | | | | | | | Stat | df | p-value |
| High | 26 | 10 | 52 | 12 | 21 | 71 | 0.48 | 0.69 | 2 | 0.71 |
| Medium | 22 | 8 | 30 | 10 | 6 | 47 | | | | |
| Low | 15 | 6 | 43 | 14 | 9 | 64 | | | | |

| Multivariate Cox regression for PFS | | | | | | |
|---|---|---|---|---|---|---|
| **Regressions** | **Coef** | **exp(coef)** | **LL CI 95% of exp(coef)** | **UL CI 95% of exp(coef)** | **se(coef)** | **p** |
| HPN Medium | -0.05 | 0.95 | 0.44 | 2.05 | 0.39 | 0.90 |
| HPN High | 0.09 | 1.10 | 0.45 | 2.68 | 0.46 | 0.84 |
| HPN Low (reference) | - | - | - | - | - | - |
| Moderately differentiated histological grade | 0.48 | 1.61 | 0.67 | 3.84 | 0.44 | 0.28 |
| Poorly differentiated histological grade | 1.24 | 3.47 | 1.48 | 8.16 | 0.44 | **0.004** |
| Well differentiated histological grade (reference) | - | - | - | - | - | - |
| ECOG 1 | -0.13 | 0.88 | 0.44 | 1.76 | 0.36 | 0.71 |
| ECOG 2 | 0.25 | 1.28 | 0.53 | 3.07 | 0.45 | 0.58 |
| ECOG 0 (reference) | - | - | - | - | - | - |
| >1 metastasis site | 1.32 | 3.73 | 1.41 | 9.84 | 0.49 | **0.008** |
| 1 metastasis site (reference) | - | - | - | - | - | - |
| Peritoneal involvement | -0.66 | 0.52 | 0.17 | 1.52 | 0.55 | 0.23 |
| No peritoneal involvement (reference) | - | - | - | - | - | - |
| Lung involvement | -1.49 | 0.22 | 0.08 | 0.60 | 0.50 | **0.003** |
| No lung involvement (reference) | - | - | - | - | - | - |
| Liver involvement | -1.47 | 0.23 | 0.08 | 0.68 | 0.55 | **0.008** |
| No liver involvement (reference) | - | - | - | - | - | - |

| Multivariate Cox regression for OS | | | | | | |
|---|---|---|---|---|---|---|
| **Regressions** | **Coef** | **exp(coef)** | **LL CI 95% of exp(coef)** | **UL CI 95% of exp(coef)** | **se(coef)** | **p** |
| HPN Medium | -0.21 | 0.81 | 0.31 | 2.13 | 0.49 | 0.67 |
| HPN High | 0.42 | 1.51 | 0.63 | 3.65 | 0.45 | 0.35 |
| HPN Low (reference) | - | - | - | - | - | - |
| Moderately differentiated histological grade | 0.72 | 2.05 | 0.92 | 4.56 | 0.41 | 0.08 |
| Poorly differentiated histological grade | 1.19 | 3.29 | 1.38 | 7.86 | 0.44 | **0.007** |
| Well differentiated histological grade (reference) | - | - | - | - | - | - |
| ECOG 1 | -0.28 | 0.76 | 0.36 | 1.60 | 0.38 | 0.47 |
| ECOG 2 | 1.02 | 2.76 | 1.07 | 7.17 | 0.49 | **0.037** |
| ECOG 0 (reference) | - | - | - | - | - | - |
| >1 metastasis site | 0.89 | 2.44 | 0.94 | 6.38 | 0.49 | 0.07 |
| 1 metastasis site (reference) | - | - | - | - | - | - |
| Peritoneal involvement | -0.21 | 0.81 | 0.29 | 2.24 | 0.52 | 0.69 |
| No peritoneal involvement (reference) | - | - | - | - | - | - |
| Lung involvement | -1.31 | 0.27 | 0.10 | 0.73 | 0.51 | **0.01** |
| No lung involvement (reference) | - | - | - | - | - | - |
| Liver involvement | -0.66 | 0.52 | 0.20 | 1.33 | 0.48 | 0.17 |
| No liver involvement (reference) | - | - | - | - | - | - |

| Multivariate Fine&Gray regression for cumulative incidence of thrombosis | | | | | | |
|---|---|---|---|---|---|---|
| **Regressions** | **Coef** | **exp(coef)** | **LL CI 95% of exp(coef)** | **UL CI 95% of exp(coef)** | **se(coef)** | **p** |
| HPN Medium | -0.58 | 0.56 | 0.18 | 1.78 | 0.59 | 0.33 |
| HPN High | -0.65 | 0.52 | 0.16 | 1.71 | 0.61 | 0.28 |
| HPN Low (reference) | - | - | - | - | - | - |
| Moderately differentiated histological grade | -0.53 | 0.59 | 0.16 | 2.21 | 0.68 | 0.43 |
| Poorly differentiated histological grade | 1.59 | 4.91 | 1.19 | 20.32 | 0.73 | **0.028** |
| Well differentiated histological grade (reference) | - | - | - | - | - | - |
| Lymphovascular invasion | -0.37 | 0.69 | 0.27 | 1.72 | 0.47 | 0.43 |
| Absent lymphovascular invasion (reference) | - | - | - | - | - | - |
| >1 metastasis site | -1.84 | 0.16 | 0.01 | 1.88 | 1.26 | 0.14 |
| 1 metastasis site (reference) | - | - | - | - | - | - |
| Peritoneal involvement | 0.78 | 2.19 | 0.22 | 21.31 | 1.16 | 0.50 |
| No peritoneal involvement (reference) | - | - | - | - | - | - |
| Lung involvement | 2.87 | 17.62 | 0.89 | 348.05 | 1.52 | 0.06 |
| No lung involvement (reference) | - | - | - | - | - | - |
| Liver involvement | 1.89 | 6.58 | 0.39 | 110.42 | 1.44 | 0.19 |
| No liver involvement (reference) | - | - | - | - | - | - |
